# Impact of Disability Certificate Requirements on Antiretroviral Initiation in HIV Care in Japan: A Retrospective Study

**DOI:** 10.64898/2026.06.24.26356015

**Authors:** Mayumi Imahashi, Tatsuya Noda, Kazumi Omata, Yoshiyuki Yokomaku, Toshibumi Taniguchi

## Abstract

**Objective:** In Japan, antiretroviral therapy (ART) for individuals living with human immunodeficiency virus (HIV) is financially supported through the Physical Disability Certification System for Immunological Impairment. However, certification requires multiple laboratory assessments after diagnosis, possibly delaying ART initiation. This study examined the impact of these eligibility requirements on ART initiation using real-world clinical data.

**Design:** Single-center retrospective cohort study.

**Setting:** Nagoya Medical Center, Japan.

**Subjects, participants:** A total of 568 patients who attended their first consultation between 2015 and 2019 were included. Of these, 434 were ART-naïve and 134 had already initiated ART at the first visit.

**Main outcome measures:** ART initiation rate, time to treatment initiation, factors associated with treatment delay, and utilization of the Physical Disability Certificate system.

**Results:** Among the 434 untreated patients, the median time to ART initiation was 42 days. Seven patients (1.6%) did not meet the Grade 4 certification requirements and remained untreated. Overall, 13 of the 568 patients (2.3%) were affected by the certification system, including those importing ART from overseas or using alternative financial support mechanisms. Non-Japanese nationality, lack of health insurance, unstable employment, and low CD4 cell counts were significantly associated with failure to initiate treatment. Among the 134 previously treated patients, 108 (80.5%) had obtained a Physical Disability Certificate.

**Conclusions:** Although relatively few patients were affected, certification requirements may delay ART initiation among socioeconomically vulnerable populations. Further multicenter and cost-effectiveness studies are needed to improve compatibility between long-term financial support systems and rapid ART initiation strategies after diagnosis.

## Introduction

In recent years, the importance of initiating antiretroviral therapy (ART) as soon as possible after HIV diagnosis has been widely recognized internationally [1]. The World Health Organization (WHO) and the Joint United Nations Program on HIV/AIDS (UNAIDS) advocates the “treat all” and “test and treat” approaches [2]. More than 90 countries have now adopted a treat-all policy [3], and rapid ART initiation, defined as starting treatment within 7 days of diagnosis, has been increasingly implemented [4]. In many of these countries, ART initiation on the day of diagnosis has become standard practice.

HIV care in Japan has also advanced considerably. As of 2020, 26,452 people living with HIV were receiving treatment, and more than 99% had achieved viral suppression [5,6]. Therefore, Japan has largely achieved the second and third UNAIDS targets. Nevertheless, some people living with HIV remain untreated after diagnosis.

In Japan, HIV treatment costs may be subsidized through the System of Medical Payment for Services and Support for Persons with Disabilities by obtaining a Physical Disability Certificate for immunological impairment. This system enables patients to receive medical expense subsidies according to their disability grade and income level, thereby facilitating long-term continuation of ART with relatively low out-of-pocket costs. However, eligibility for the certificate requires multiple laboratory test results obtained at specified intervals after HIV diagnosis. The extent to which these requirements may delay ART initiation has not been sufficiently investigated.

To date, few studies in Japan have examined the impact of Physical Disability Certificate eligibility requirements on the timing of ART initiation using real-world clinical data. In this study, we investigated the effect of the Physical Disability Certificate system on ART initiation. We hypothesized that the certification requirements may delay ART initiation among socioeconomically vulnerable patients. Specifically, we examined (1) treatment initiation rates and factors associated with delayed treatment among patients who had no initiated ART at their first hospital and (2) the actual utilization of the certificate system among patients who had already initiated ART at the time of their first visit.

## Methods

### Patients

This study included patients who attended their first consultation at Nagoya Medical Center between 2015 and 2019. Because all eligible patients were enrolled, a formal a priori sample size calculation was not performed. For all patients at their initial visit, the following data were extracted from medical records: date of first visit, sex, age at first visit, nationality, ART status, and possession of a Physical Disability Certificate.

For patients who had not initiated ART at the time of their first visit, the following baseline characteristics were additionally collected from medical records: health insurance status, employment status, CD4 cell count at the first visit, HIV-RNA viral load, disease stage at presentation, presence of acute infection, ART initiation status by December 2021, date of treatment initiation, municipality of residence, and, for untreated patients, the reason for remaining untreated.

For patients who had already initiated ART at the time of their first visit, the following additional baseline data were collected: use of the System of Medical Payment for Services and Support for Persons with Disabilities, reason for not applying for a Physical Disability Certificate, whether HIV had been diagnosed overseas, and the method of obtaining ART as of December 2021.

### Physical Disability Certificate System

In Japan, medical expense subsidies for HIV treatment are provided on the basis of certification for immunological impairment through the Physical Disability Certificate system. Certificates are classified into Grades 1–4 according to the severity of immunological impairment. Based on the disability grade and the patient’s income category, patients may receive financial support through the System of Medical Payment for Services and Support for Persons with Disabilities, as well as additional subsidies provided by local governments.

Application for a Physical Disability Certificate requires repeated confirmation of HIV-related laboratory parameters, including CD4 cell count and HIV RNA levels, at specified intervals after diagnosis. Following certification, a medical certificate is issued by the local government. Details regarding the classification of immunological impairment and certification requirements are presented in Table 1.

**Table 1.**
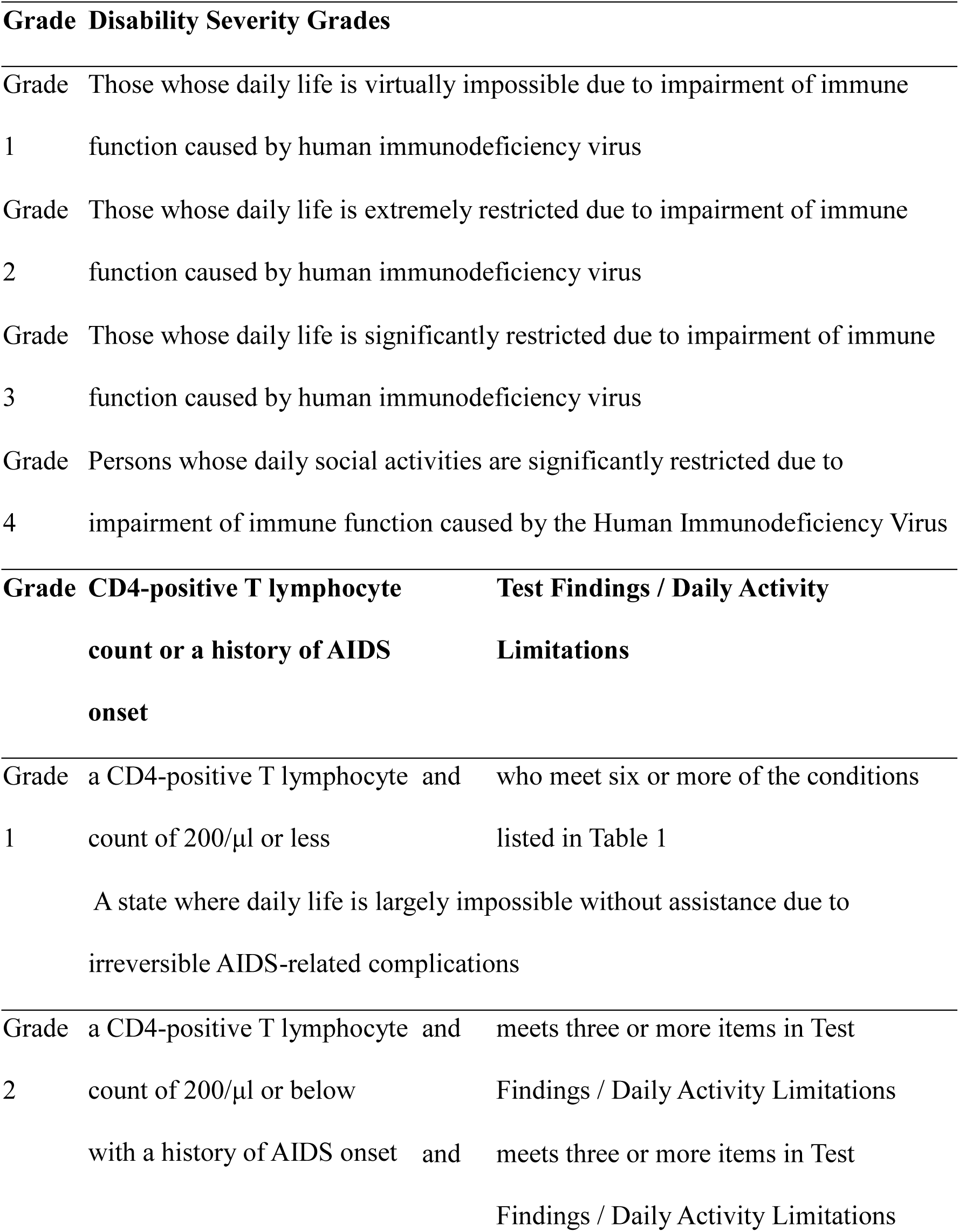
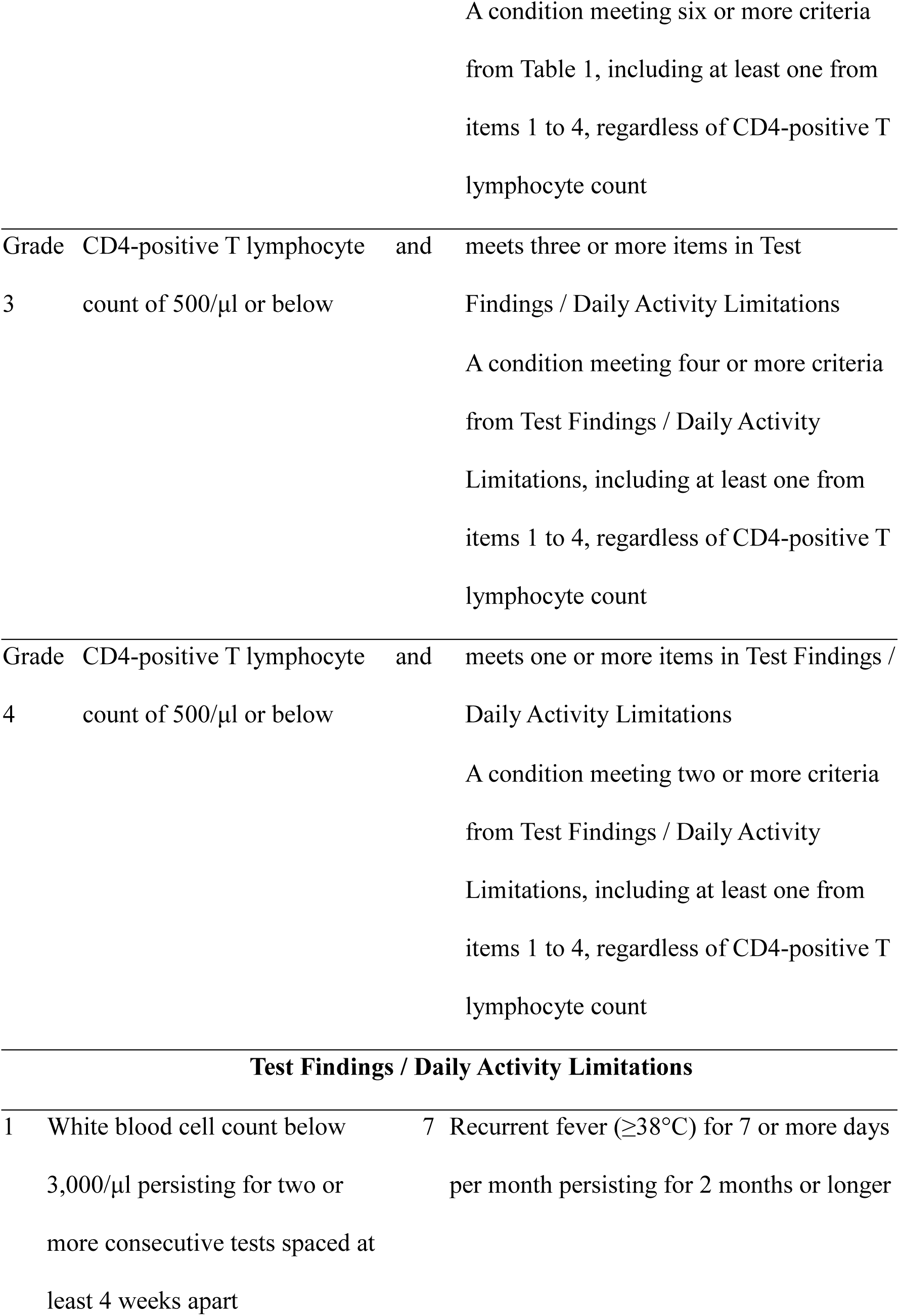
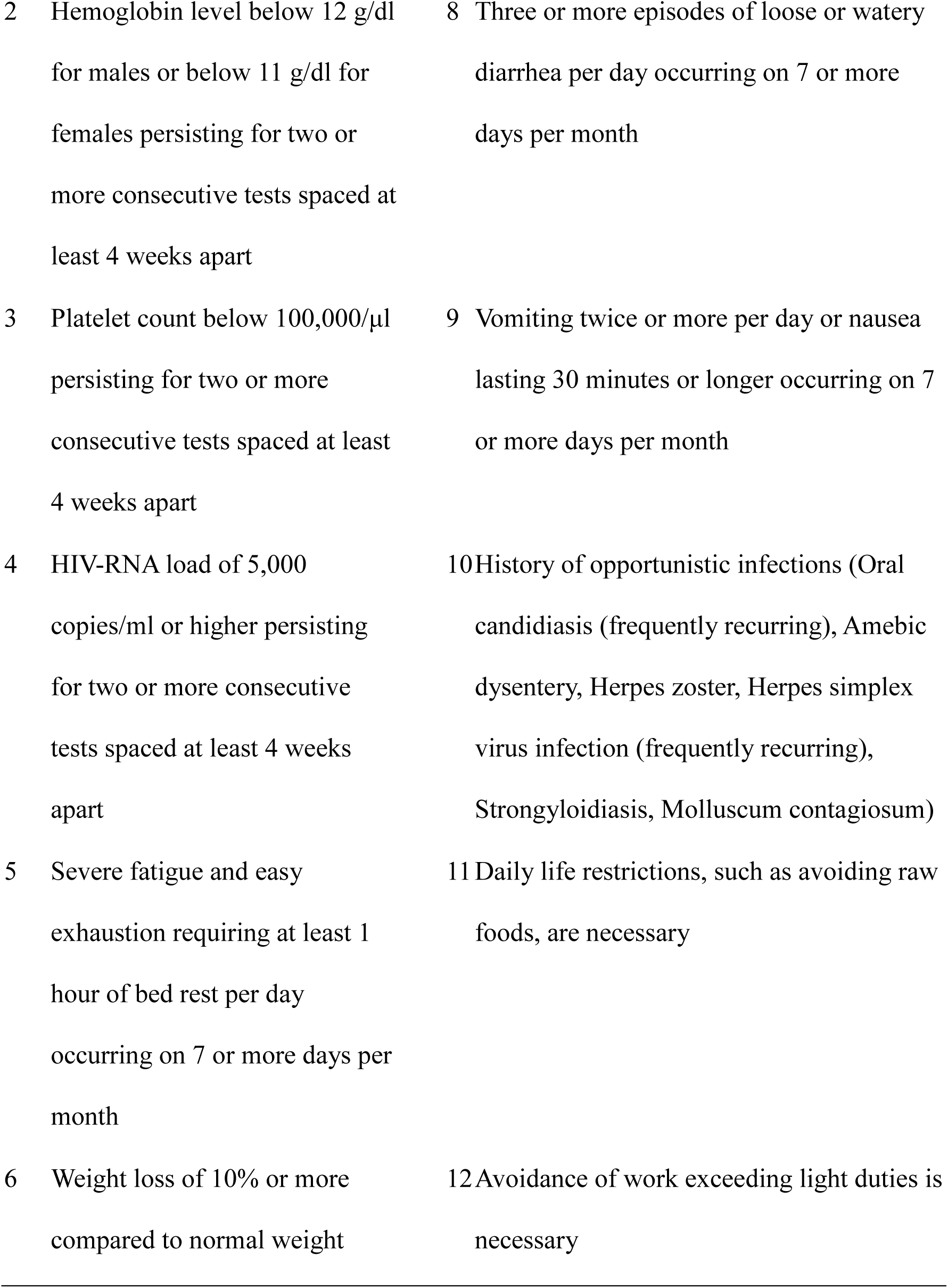
Classification criteria for Physical Disability Certification due to HIV-related immunological impairment in Japan.

### Statistical Analysis

Continuous variables were analyzed using the Mann-Whitney U test, whereas categorical variables were analyzed using the chi-square test. Among patients who had not initiated ART at their first hospital visit, multivariate analyses were performed to identify factors associated with treatment status as of December 2021 and time to ART initiation. Variables included in the multivariate models were selected a priori based on clinical relevance and findings from prior studies.

Statistical significance was defined as p < 0.05. All statistical analyses were performed using Stata software (ver. 15.0). To evaluate differences among municipalities in the time required to obtain physical disability certification, residential addresses of untreated patients who were not diagnosed during the AIDS stage were mapped, and spatial autocorrelation analysis was conducted. ArcGIS (ver. 10.8) was used for the spatial analysis. Statistical significance was defined as p < 0.05.

### Ethics Statement

This study was approved by the Clinical Research Ethics Committee of the National Hospital Organization Nagoya Medical Center (Approval number: 2021-079). As this was a retrospective observational study, the requirement for informed consent was waived in accordance with applicable ethical guidelines.

## Results

Between 2015 and 2019, 568 patients attended the Department of Infectious Diseases at Nagoya Medical Center for their first consultation (Figure 1). Of these, 434 were ART-naïve at the time of the first visit, while 134 had already initiated ART. Among the 434 ART-naïve patients, 33 remained untreated or had an unknown treatment status as of December 2021. Of the 134 patients already on ART at the time of their first visit, 108 (80.5%) had obtained a Disability Certificate.

**Figure 1.**
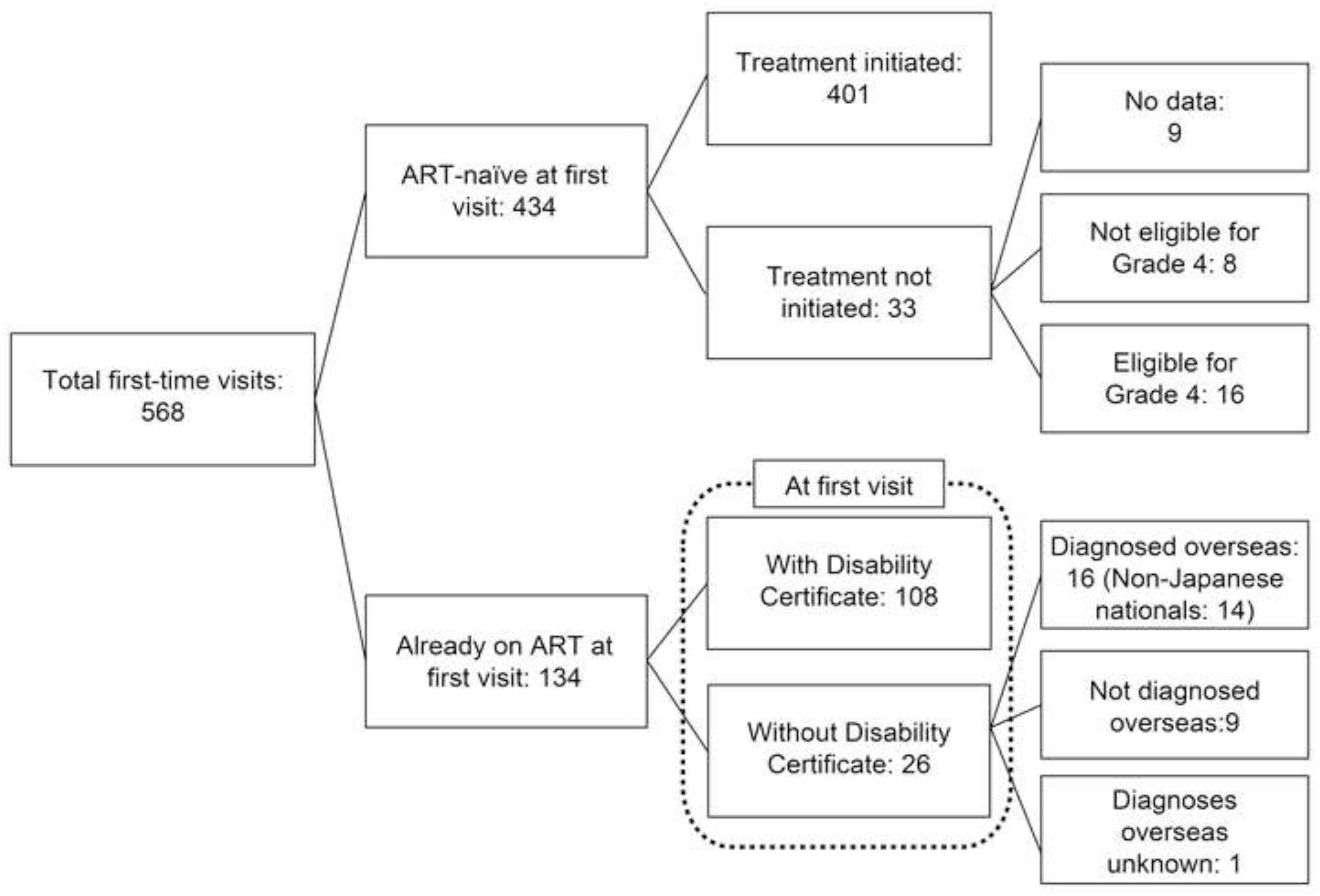
Patient flow at first visit to Nagoya Medical Center

### Patients Untreated at the First Visit

#### 1) Patient characteristics (Table 2A)

No significant differences were observed between patients who initiated ART and those who did not (as of December 2021) in terms of sex, disease stage at first visit, or presence of acute HIV infection. In contrast, significant differences were observed with respect to age, nationality (Japanese vs non-Japanese), health insurance status, employment status, CD4 cell count at the first visit, and HIV RNA viral load at the first visit.

**Table 2A.**
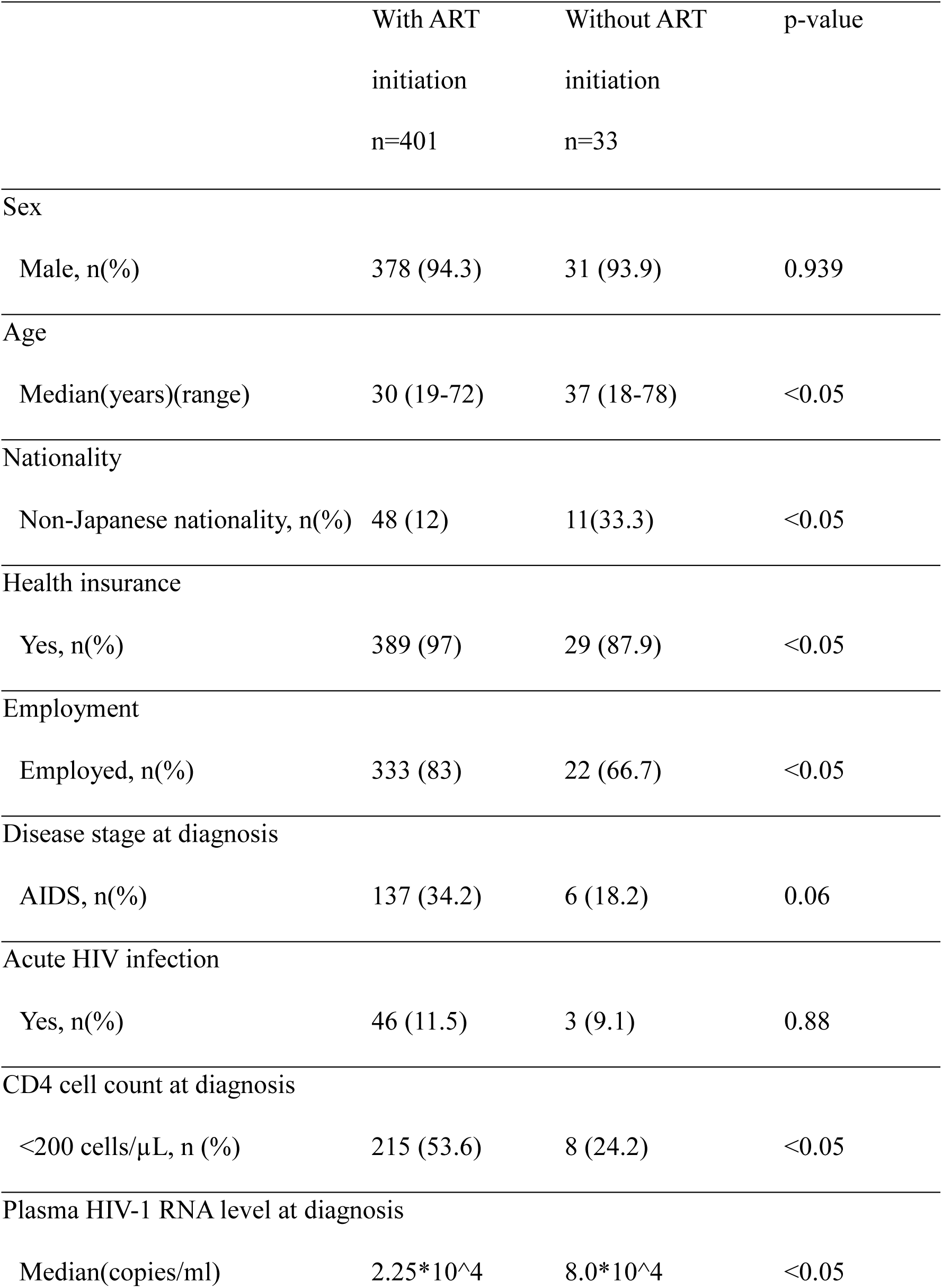
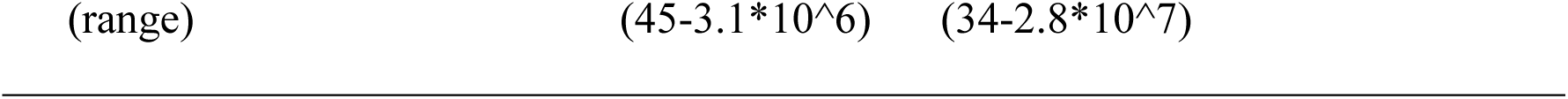
Baseline characteristics of treatment naïve patients with and without ART initiation.

#### 2) Days until treatment initiation (Figure 2)

Time to ART initiation is shown in 30-day intervals. The median time to treatment initiation was 42 days, with the most common time being within 0-30 days (31 patients). Ninety percent of patients who initiated treatment started therapy within 161 days of their first visit. The median time to treatment initiation by disease stage was 14 days for those diagnosed with AIDS and 70 days for those diagnosed with non-AIDS conditions.

**Figure 2.**
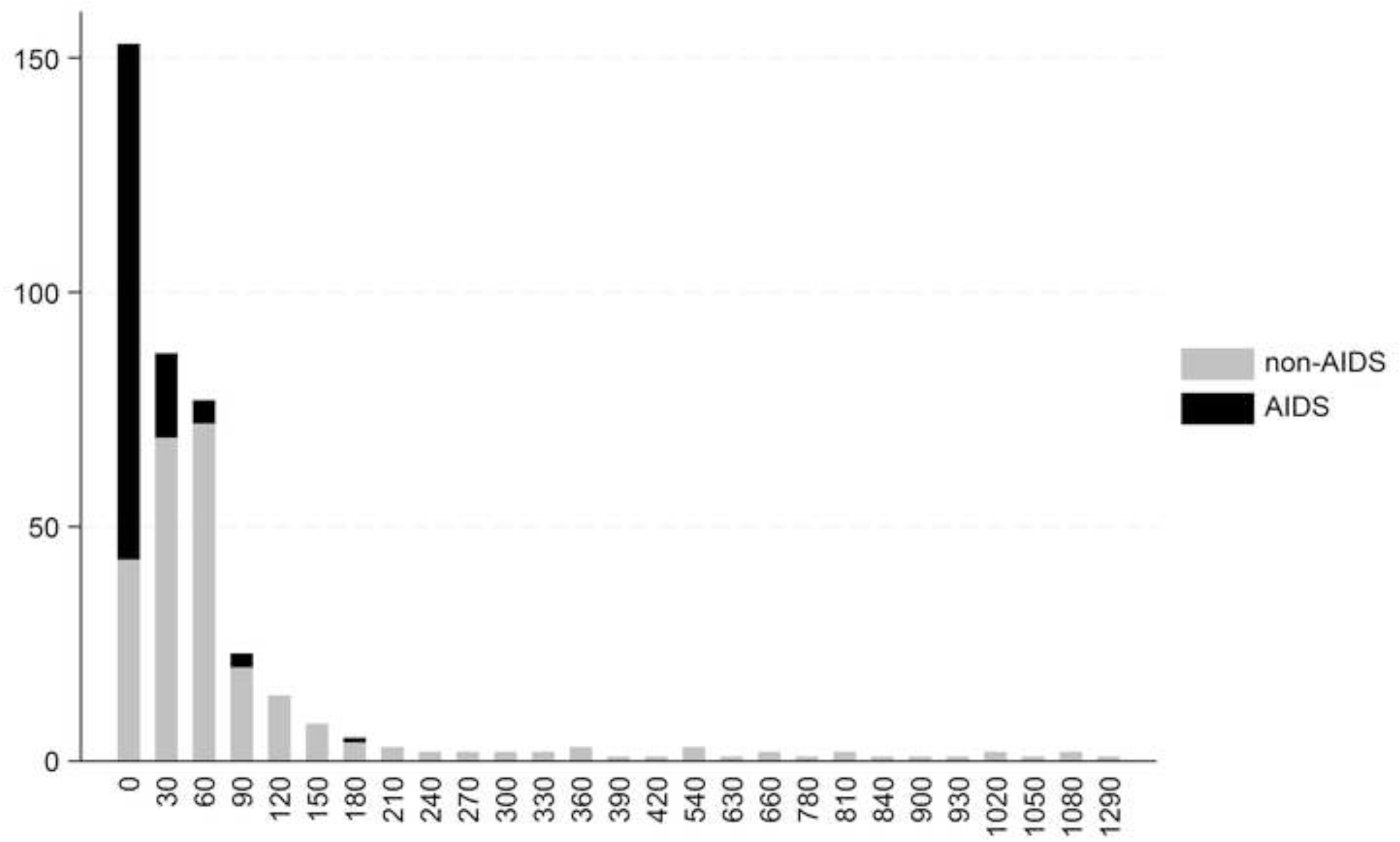
Time to treatment initiation

#### 3) Reasons for non-initiation of treatment (Figure 3A)

Among the 33 untreated patients, data were not available for nine patients (27.2%) due to a discontinuation of visits or transfer to another hospital. Seven patients (21.2%) were unable to obtain a Grade 4 Physical Disability Certificate, and 17 (51.5%) were eligible for Grade 4 but had not initiated treatment. Among the seven patients who were not eligible for Grade 4 certification, none met the Grade 4 eligibility criteria based on either viral load or CD4 count. Among the 17 patients who were eligible for Grade 4 but did not receive treatment, 11 were waiting to meet the criteria for Grade 3 disease. Among those 11 patients waiting for Grade 3 eligibility, five were still under follow up as of December 2021, three had been transferred to another hospital, and three had discontinued visits. Overall, among the 434 ART-naïve patients, seven (1.6%) did not meet the requirements for Grade 4 certification.

**Figure 3.**
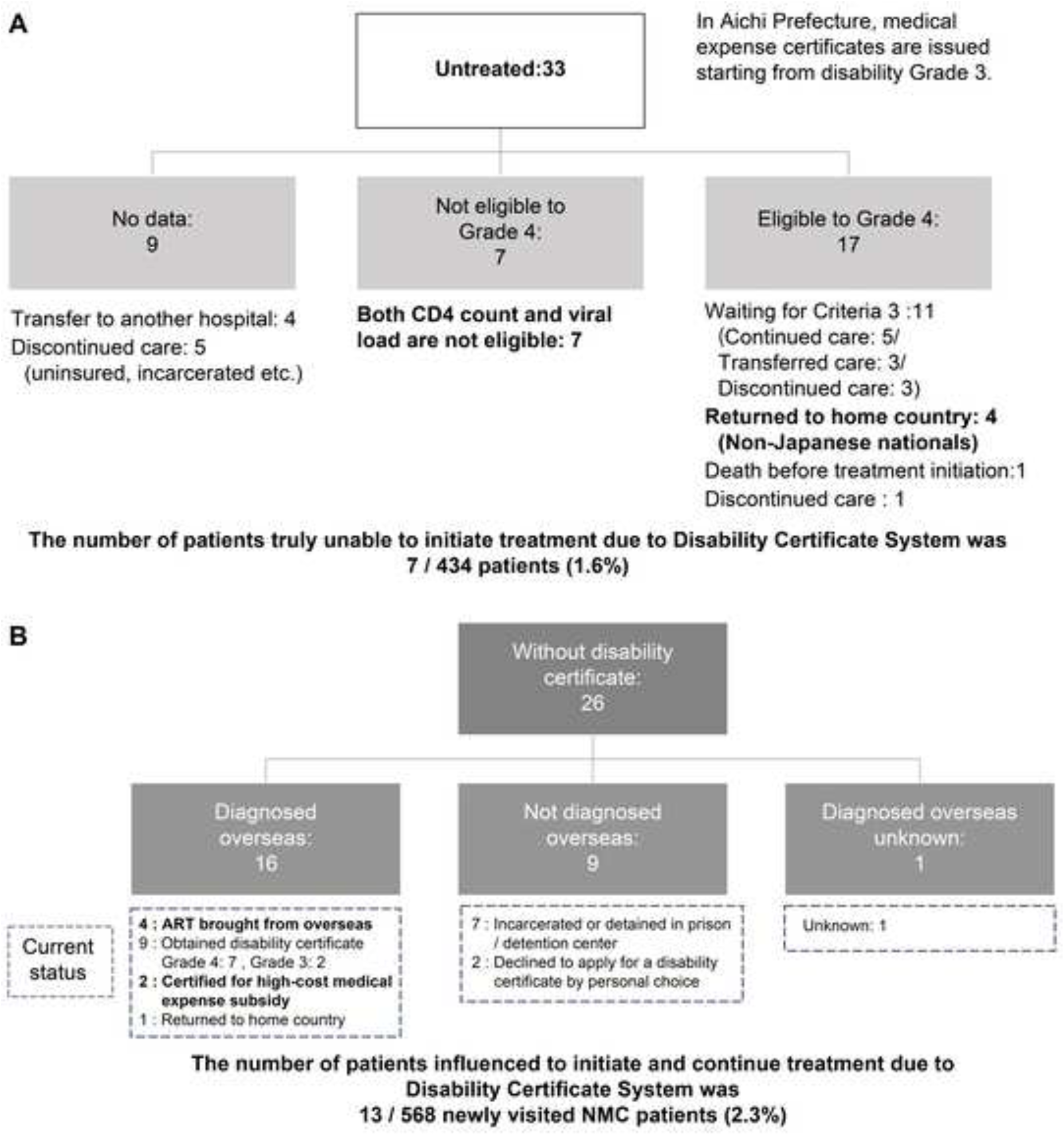
Reasons for not accessing available medical and social support among study participants: **(A)** Antiviral therapy and **(B)** Disability certificate.

#### 4) Factors associated with treatment initiation (Supplementary Table 1A)

The factors associated with treatment initiation were nationality, health insurance status, employment status, and CD4 count at the first visit. For nationality, patients of non-Japanese nationality had an odds ratio of 0.21 for treatment initiation compared to patients of Japanese nationality.

#### 5) Factors associated with time to treatment initiation (Supplementary Table 1B)

Sex and CD4 cell count at diagnosis were significantly associated with time to ART initiation. Female patients had a shorter time to treatment initiation compared with male patients.

#### 6) Patient residence (registered address) and treatment days (Supplementary Figure 1)

Patients diagnosed at non-AIDS status were divided into two groups based on the median time to ART initiation (70 days). Residential locations were mapped, and spatial autocorrelation analysis was performed. The Moran’s I Index was 0.14 (p = 0.76), indicating no spatial clustering in time to treatment initiation by municipality of residence.

#### 7) Treatment continuity

Among the 401 patients who were ART-naïve at first visit and subsequently initiated treatment, treatment continuation rates were 97.1% at 1 year, 95.6% at 3 years, and 93.1% at 5 years after initiation (as of December 2021).

#### Patients Already on Treatment at First Visit

Between 2015 and 2019, 134 patients attended their first consultation after initiation ART. At the time of their first visit, 108 patients (80.5%) had obtained a Physical Disability Certificate, while 26 (19.4%) had not.

#### 1) Patient characteristics (Table 2B)

Among patients already on ART at first visit, the median age was 40 years (range: 21-67 years), and 121 patients (90.3%) were male. Ninety-nine patients (73.9%) were Japanese. No significant differences were observed in Physical Disability Certificate acquisition by age or sex (age: p = 0.23; sex: p = 0.131). However, significant differences were observed according to nationality, health insurance status, and employment status.

**Table 2B.** Baseline characteristics of treated patients with and without disability certificate.

|  | With Disability<br>certificate<br>n=108 | Without Disability<br>certificate<br>n=26 | p-value |
| --- | --- | --- | --- |
| Sex |  |  |  |
| Male n(%) | 100(92.6) | 21(80.8) | 0.131 |
| Age |  |  |  |
| Median(years)(range) | 38 (22-57) | 41 (21-67) | 0.23 |
| Nationality |  |  |  |
| Non-Japanese nationality, n(%) | 19 (17.6) | 16 (61.5) | <0.05 |

#### 2) Characteristics of patients without a Physical Disability Certificate (Figure 3B)

Among the 26 patients without a Physical Disability Certificate, 16 (61.5%) had been diagnosed with HIV overseas, nine (34.6%) had been diagnosed in Japan, and one case (3.8%) was unknown.

Among the 16 patients diagnosed overseas, nine subsequently obtained diagnostic documentation from the country of diagnosis; of these, seven obtained a Grade 4 certificate and two obtained a Grade 3 certificate. Four patients had obtained ART abroad. Two patients received treatment under the high-cost medical expense ceiling system, and both were classified as “Category E.” The remaining patients returned to their home country.

Among the nine patients diagnosed in Japan, seven did not obtain a Physical Disability Certificate because they were incarcerated or detained. The remaining two declined to apply for the certificate (Figure 3B).

Overall, seven patients were ineligible for Grade 4 certification at the first visit. In total, 13 of 568 patients (2.3%) were affected by system-related barriers to obtaining a Physical Disability Certificate, including Grade 4 ineligibility (n = 7), ART obtained overseas (n = 4), and use of the high-cost medical expense ceiling system (n = 2) (Figure 1).

## Discussion

In this study, 2.3% of all patients were unable to benefit from the system at their first visit. This included patients who could not initiate ART because they did not obtain a Physical Disability Certificate, as well as those who continued treatment by importing ART from overseas or receiving care under the high-cost medical expense ceiling certification. Among patients who were ART-naïve at the first visit, 1.6% were unable to initiate treatment due to the eligibility requirements for Physical Disability Certificate.

In many countries, rapid ART has become the standard approach of care, with treatment initiated on the day of diagnosis or within seven days [3]. This approach is supported by the START trial, which demonstrated that immediate ART initiation in asymptomatic individuals with CD4 counts > 500 cells/mm^3^ significantly reduces the risk of serious AIDS-related events, serious non-AIDS-related events, and death compared with deferred treatment initiation [1]. Rapid ART is also associated with earlier viral suppression and improved treatment retention [7–9]. In countries where rapid ART is implemented, there are mechanisms to cover the initial treatment costs, such as temporary starter packs [10], bridge funding through public funds [11], and emergency ART that can be used even without insurance [12]. In these systems, subsidies for medical expenses are not prerequisites for treatment initiation. This suggests that the association between treatment initiation and medical assistance costs differs between Japan and other countries.

In Japan, the Physical Disability Certification system was introduced to reduce the financial burden of long-term HIV treatment. While the system is effective in limiting out-of-pocket costs over time, certification requires laboratory assessments at specified intervals prior to approval. Consequently, eligibility confirmation may take time, and some required criteria may become less applicable once treatment has already been initiated. This structure may therefore create an implicit expectation that certification procedures should be completed before ART initiation.

Although the proportion of patients affected by the system was small in this study, such an effect is unlikely to affect all patients equally. In fact, patients who did not meet the certification requirements or who were unable to use the system were foreign nationals and patients with unstable health insurance or employment statuses. This trend is consistent with that reported by Hashiba et al. [13]. They concluded that the 5-year attendance rate among foreign-born patients was 75.5%, whereas that among Japanese patients was 94.1%, which was statistically significant (p<0.001). This study suggests that patients in socioeconomically vulnerable positions are more likely to be affected by system constraints.

Furthermore, these institutional effects are likely to become apparent not during the long-term continuation phase of ART but during the early phase from diagnosis to treatment initiation. This period is also one in which discontinuation of visits or transfer to other institutions is more likely to occur. As such, clearly explaining the process leading to treatment initiation and supporting continued commitment to follow-up visits are clinically important.

The Physical Disability Certificate system was originally designed to support the continuation of treatment. Long-term continuation of ART in Japan involves practical challenges that underscore the importance of sustained support systems. A nationwide analysis of the National Database (NDB) demonstrated that approximately 19% of individuals living with switched their anchor drug between first and second ART regimens, with higher long-term switch rates observed for NNRTI- and PI-based regimens, while INSTI-based regimens demonstrated lower switch rates over an 8-year period [14]. Furthermore, a nationwide cohort study using the same database reported that 81.5% of 28,089 individuals had at least one chronic comorbidity, with multimorbidity and polypharmacy increasing with age [15]. These findings underscore that long-term HIV management requires not only sustained access to ART but also ongoing care for comorbid conditions and regimen optimization, reinforcing the importance of financial support systems such as the Physical Disability Certificate. However, owing to its certification requirements, a temporal gap may arise between the system and the rapid initiation of treatment immediately post diagnosis. Consequently, while the system itself plays an important role in reducing the burden of medical expenses, it may inadvertently become a factor that delay treatment initiation from the perspective of international treatment strategies aimed at treatment immediately post diagnosis. This misalignment between institutional design and clinical reality is not unique to Japan. Grobman et al. highlighted similar issues in life and disability insurance systems, where coverage decisions may still be influenced by HIV status, despite substantial improvements in prognosis due to modern ART, illustrating a broader lag between policy frameworks and advances in HIV care [16].

Accordingly, these findings do not question the effectiveness of the Physical Disability Certificate system. Rather, they highlight a structural and temporal gap between its strengths in supporting long-term treatment and its alignment with strategies emphasizing immediate ART initiation after diagnosis.

Overall, the results suggest that while the Physical Disability Certificate system plays an important role in sustaining long-term ART, its relationship with the timing of treatment initiation warrants further consideration. Future research should include simulation studies and cost-effectiveness analyses to evaluate models that support immediate ART initiation after diagnosis while maintaining financial sustainability.

### Limitations

This study has several limitations. First, it was a single-center retrospective study of patients attending Nagoya Medical Center; therefore, generalizability to other regions may be limited. This is particularly relevant in HIV care, where welfare and administrative systems may vary across municipalities and potentially influence treatment initiation. Second, socioeconomic status could not be fully captured. In this study, socioeconomic factors were approximated using employment and health insurance status. However, socioeconomic factors may also include residency status, housing stability, Japanese language proficiency, health literacy regarding medical and welfare systems, and experiences of stigma, which were not available. Third, the differences in system operations among municipalities could not be fully reflected in this study. Although the physical disability certification system is nationwide, its operations differ among municipalities. For instance, there may be differences in whether the physical disability certificate and the system of medical payment for services and support for individuals with disabilities can be applied simultaneously, the number of days required from application to issuance, and the level of understanding of the HIV-related system among staff members at service counters. Additionally, because there are no data that quantify the operation of the system by municipality and because there is a small possibility that the municipality where the application was submitted differed from the current residential address, these factors cannot be completely ruled out. Therefore, several directions for future research are proposed. First, a prospective multicenter cohort study involving HIV treatment facilities across multiple prefectures with different welfare systems should be conducted to assess the generalizability of these findings. Second, future studies should incorporate more comprehensive measures of socioeconomic status, including residency status, residential stability, Japanese language proficiency, health literacy regarding the medical and welfare systems, and experiences of social stigma, potentially through validated questionnaires administered at the time of initial consultation. Third, a quantitative survey of municipal-level differences in system operations would be valuable. Collaborating with local governments to systematically document procedural variations could provide the data necessary to include municipal-level factors as covariates in a multilevel analysis.

## Conclusions

In this single-center retrospective analysis, 2.3% of all first-visit patients were affected in terms of treatment initiation or continuation because of difficulty in accessing or utilizing the current eligibility requirements for the Physical Disability Certificate system. Among patients who were ART-naïve at the first visit, certification requirements may have functioned as a barrier to ART initiating in 1.6% of cases. Although the proportion of affected patients was small, the impact appears to be concentrated among socioeconomically vulnerable individuals during the early phase following HIV diagnosis. In clinical practice, it is important to clearly explain the pathway to treatment initiation and to provide support that facilitates continuous engagement in care. Future research should include simulation and cost-effectiveness analyses of immediate ART initiation after diagnosis. Such studies are needed to evaluate how to preserve the strengths of the system in supporting long-term treatment continuity while ensuring compatibility with rapid treatment initiation strategies. For instance, comparing estimated out-of-pocket costs under the high-cost medical expense benefit system without a Physical Disability Certificate verses costs with certification, using publicly available Japanese healthcare expenditure data, would help quantify the financial implications of current eligibility requirements and inform policy discussions on alternative early-phase support mechanisms.

## Data Availability

The data that support the findings of this study are available on request from the corresponding author. The data are not publicly available due to privacy or ethical restrictions.

## Acknowledgements

Editorial support, including medical writing, table assembly, and high-resolution image creation, was provided by Editage and Cactus Communications. The authors would like to thank all the participants of the study.

